# Development of a multi-scanner facility for data acquisition for digital pathology artificial intelligence

**DOI:** 10.1101/2023.11.07.23297408

**Authors:** Matthew P. Humphries, Danny Kaye, Gaby Stankeviciute, Jacob Halliwell, Alexander I Wright, Daljeet Bansal, David Brettle, Darren Treanor

**Affiliations:** National Pathology Imaging Cooperative, Leeds Teaching Hospitals NHS Trust, Leeds, UK; University of Leeds, Leeds, UK; Department of Histopathology, Leeds Teaching Hospitals NHS Trust, Leeds, UK; Department of Clinical Pathology and Department of Clinical and Experimental Medicine, Linköping University, Linköping, Sweden; Centre for Medical Image Science and Visualization (CMIV), Linköping University, Linköping, Sweden

**Keywords:** Digital pathology, slide scanning, whole slide imaging, whole slide scanners, multi-scanner facility, Artificial Intelligence

## Abstract

Whole slide imaging (WSI) of pathology glass slides with high-resolution scanners has enabled the large-scale application of artificial intelligence (AI) in pathology, to support the detection and diagnosis of disease, potentially increasing efficiency and accuracy in tissue diagnosis.

Despite the promise of AI, it has limitations. “Brittleness” or sensitivity to variation in inputs necessitates that large amounts of data are used for training. AI is often trained on data from different scanners but not usually by replicating the same slide across scanners. The utilisation of multiple WSI instruments to produce digital replicas of the same glass slides will make more comprehensive datasets and may improve the robustness and generalisability of AI algorithms as well as reduce the overall data requirements of AI training.

To this end, the National Pathology Imagine Cooperative (NPIC) has built the AI FORGE (**F**acilitating **O**pportunities for **R**obust **G**eneralisable data **E**mulation), a unique multi-scanner facility embedded in a clinical site in the NHS to (a) compare scanner performance and (b) replicate digital pathology image datasets across WSI systems.

The NPIC AI FORGE currently comprises 15 scanners from 9 manufacturers. It can generate approximately 4000 WSI images per day (approximately 7Tb of image data). This paper describes the process followed to plan and build such a facility.

## Introduction/Background

The adoption of digital pathology is growing rapidly, with increasing digitisation of routine clinical practice. In parallel, advances in computational pathology and artificial intelligence (AI), are increasingly being seen in pathology, driven by academic and industry efforts.

To ensure AI is robust, researchers use a variety of techniques in the development of AI tools, such as: collating images from varied institutions, using different H&E staining protocols and tissue types, and using diverse pathologist annotation and slide label inputs. However few studies consider the landscape of whole slide imaging (WSI) platforms available on the market and the impact this may have on algorithm accuracy. There is a diverse range of digital pathology vendors providing WSI platform solutions, each system capable of producing high resolution images. A comprehensive review of WSI imaging hardware available was presented by Patel *et al*. whereby the wide range of cameras and sensors, slide loading and handling, objectives and magnifications, scanning and focus methods, as well as physical parameters such as size and weight, is summarised [1].

From this selection, laboratories may struggle to choose systems that meet their individual needs. Though the myriad of scanning platforms is daunting for a laboratory starting their digital adoption journey, the diversity of platforms available also presents an important opportunity to understand their impact on the diagnostic process and laboratory workflow.

We have investigated factors which may impact diagnostic pathology in digital pathology and AI, such as colour reproduction [2-4], histological and scanning quality control [5-8], and have used this to produce practical information on clinical deployment [9].

As clinical use of AI emerges, AI tools have demonstrated impressive accuracy in isolation and on test datasets but can suffer from a reduction in performance when presented with real world cases from different institutions and different scanner instruments [10]. Introducing image training data from multiple scanner vendors may have the potential to provide the robust training required to create more generalisable AI. However, provision of broader additional image data simply by accessing data from different institutions using different scanner platforms is not a systematic or complete solution. In fact, a strategy wholly reliant on combining datasets from different institutions who use different scanner instruments could lead to additional bias due to differences in case mix and pre-digitisation sample preparation.

The appreciation of inter-scanner variability is a key consideration in the development and evaluation of AI tools. Identifying the variation in images from different scanner vendors that influence AI algorithmic decision making will enable AI tools to train their output decisions across images from multiple sources and reduce the potential for batch effects.

A national digital pathology program, National Pathology Imaging Co-operative (NPIC) - (https://npic.ac.uk/), will digitise over 40 NHS hospitals in a single national network to support the clinical adoption of digital pathology and the development and use of AI. An important consideration in such a program is how to ensure AI is robust, and how it may perform in the real world using different scanner models and instruments. AI trained on data replicated on multiple instruments may produce more robust AI and reduce the overall data requirements for AI development (as replicas of the same image may have more training value than adding different images from different sites on different scanners).

To explore this training capability, NPIC has created the AI FORGE (**F**acilitating **O**pportunities for **R**obust **G**eneralisable data **E**mulation), a unique multi-scanner facility to replicate digital pathology image datasets on multiple research and clinical systems, embedded within an NHS hospital. The aims of the FORGE are to (a) compare scanner performance and (b) replicate digital pathology image datasets across WSI systems. This article describes the planning and work involved in developing the AI FORGE, comprising 15 scanner instruments from 9 vendors.

## Methodology/Approach

### Facility planning

The initial step in establishing the AI FORGE was to secure dedicated laboratory space suitable for a substantial new digital pathology facility. A project planning team was convened to consider all aspects of the installation including, staffing, building and scanner room security access, sufficient space for the receipt and preparation of slides, short- and long-term storage of slides, appropriate benching, suitable PC workstations, air-conditioning, and an optimised workflow arrangement to allow efficient daily operation when scanning across multiple systems. Figure 1 shows the facility plan.

**Figure 1.**
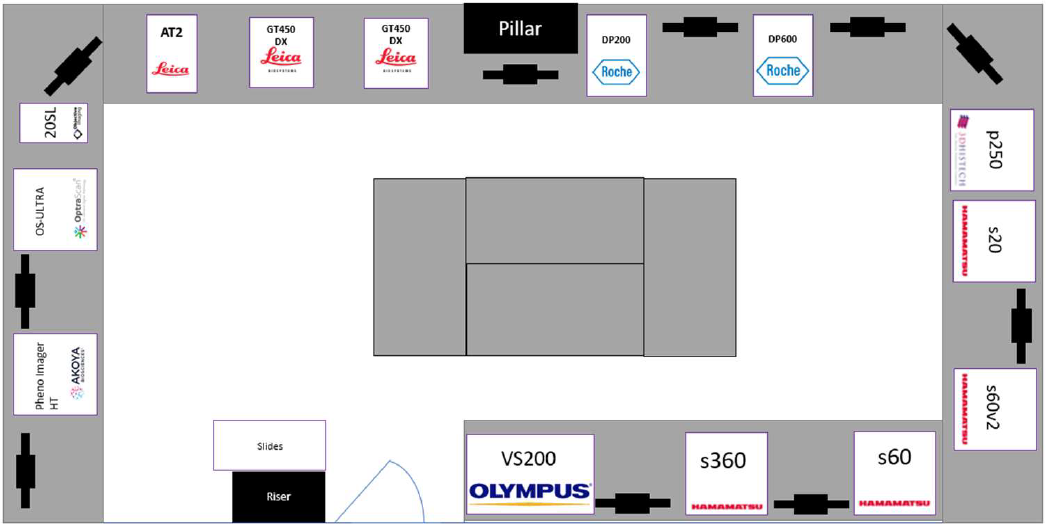
FORGE facility plan prior to install.

Further to the logistical and environmental deployment of the facility on-site, technical requirements were evaluated and factored into the centre design including sufficient electrical power supply, equipment and PC networking, data storage and the compute required for data analysis. A research image management system (RIMS) where all images would be ingested from each scanner was also required. The detailed requirements are reported below.

### Procurement

Large procurements of clinical and information technology systems are often complex and multi-staged processes. To expedite the facility establishment, we created a streamlined approach to digital pathology systems procurement. In parallel with the physical space customisation was the formation of an NHS digital pathology procurement framework agreement for the acquisition of clinical grade high-throughput digital pathology equipment, accessible to all UK NHS Trusts and NPIC partners.

We created the Digital Pathology Solutions Framework [11], and published an invitation to tender, administered by Health Trust Europe (https://www.healthtrusteurope.com/). Following internal review and vendor acceptance, the framework was established and became live in May 2021, at which point procurement activity began following the NPIC Health Trust Europe Digital Pathology Solutions Framework roadmap shown in figure 2. Briefly, the roadmap begins with the completion of a digital access form, followed by confirmation of funding by the NPIC board. Meetings are set to outline specifications of the equipment required and supplier(s) are engaged. A quotation is raised following product demonstration(s), if required. A call-off contract is established between all parties, followed by contract review, and signature. A purchase order is raised, and systems are deployed and installed as per the agreed terms.

**Figure 2.**
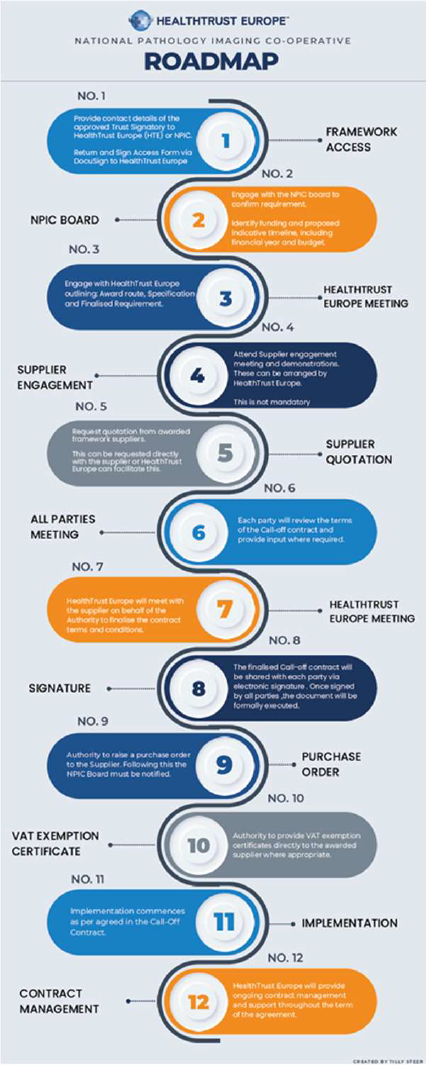
NPIC Health Trust Europe Digital Pathology Solutions Framework road map.

Over a 6-month period, NPIC acquired 15 scanners from 9 vendors. These include: Two Leica Biosystems GT450s and a Leica Biosystems AT2; one Roche DP200; one Roche DP600; four Hamamatsu scanners (S360MD, S60, S60v2MD, S20); one 3DHistech P250 Flash; one Olympus VS200; one Objective Imaging Glissando; one Akoya Bioscience PhenoImager; one OptraScan - OS-Ultra; and one Grundium Ocus.

## Results

### Dedicated FORGE team

In parallel to the project planning was the requirement for dedicated staff with the skills and expertise to deliver the scanning activity of the AI FORGE. An operations manager and project coordinator were appointed to oversee the delivery of the facility and recruit a specific team of highly skilled staff. A team of six Digital Pathology Scientists and one Research Technician were recruited. The team comprises several senior and junior NHS trained Biomedical Scientists with experience of diagnostic histopathology, as well as research active laboratory scientists with published research in the field of computational pathology. The team is supported by a wider team of NPIC innovation project managers, a business manager, and project support officers as required by the needs of the organisational workload. In total the NPIC FORGE team comprises eight full time equivalent staff to operate, with a further supporting team of seven technical, project, and operational managers.

### Scanners

WSI scanners were selected from the framework with the aim of comprehensive cover of the range of available systems on the market. The makes, models, capacity, size/weight, loading method, microns per pixel, file type, and regulatory status information for the systems selected for installation within the AI FORGE are detailed in table 1.

**Table 1.**
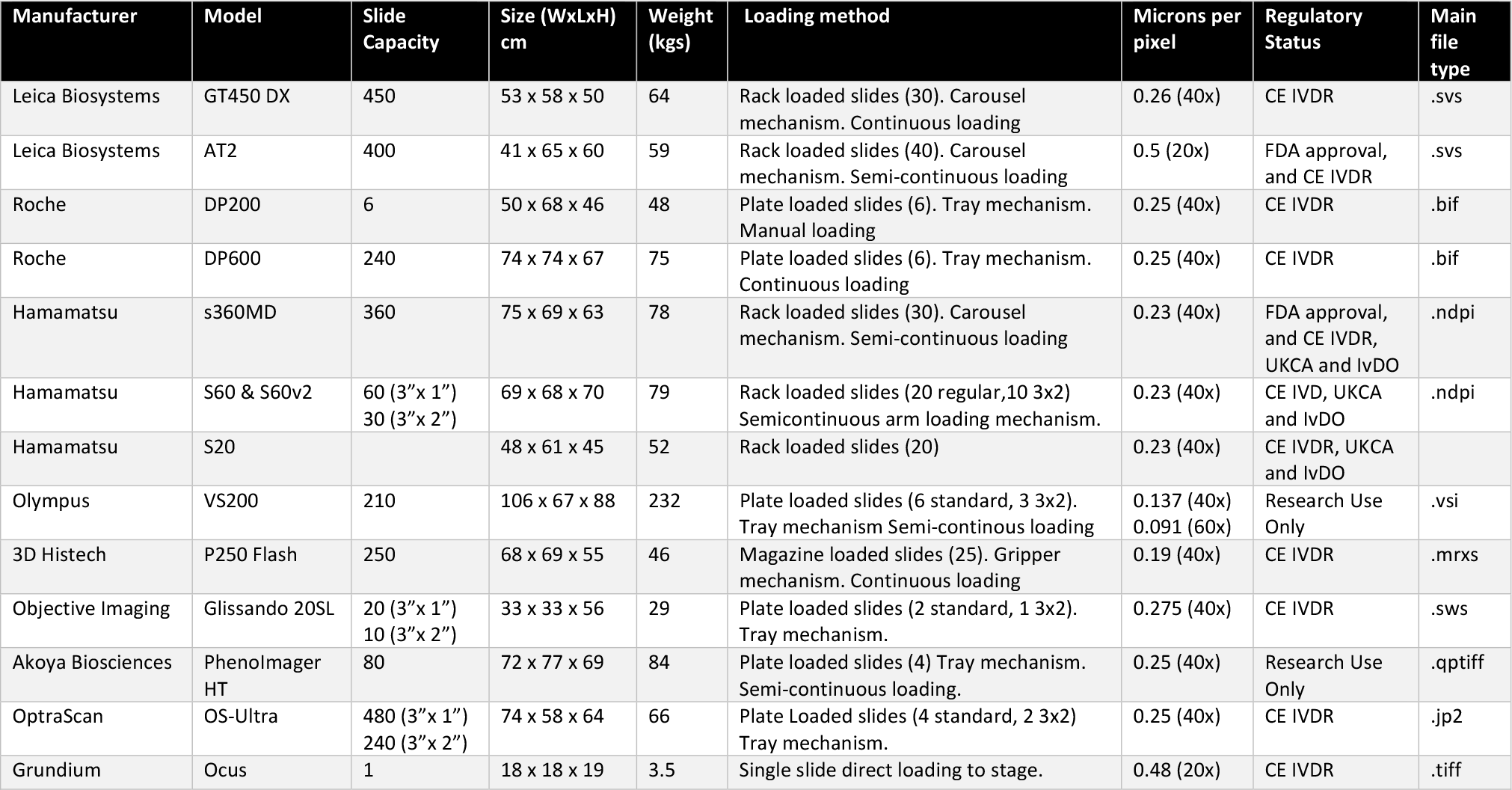
WSI systems in the NPIC AI FORGE detailing manufacturer, model, capacity, size/weight, loading method, microns per pixel, file type, and regulatory status.

The capacity and capabilities of the AI FORGE for WSI include batch slide scanning from as few as 6 slides (e.g. a single load of a Roche DP200), to a batch of 450 slides (the single loading capacity of a Leica GT450). WSI can be captured in multiple image magnifications, from 10x to 60x (the latter which requires automatic oil dispensing on the Olympus VS200). The most common magnification in the scanning facility is 40x (approximately 0.25 microns per pixel).

All 15 scanning platforms are capable of scanning 3”x1” slides, with 6 of these capable of scanning 3”x2” slides. Figure 3 displays the replication of a single 3”x2” slide on the 6 platforms capable of scanning these slides, with a single 3”x1” slide replicated on the remaining 8 WSI platforms. Scanned using default manufacturer settings, the images pixel dimensions range from 0.8 to 61 gigapixels, and image file sizes range from 1.47 to 6.51Gb (19.3 to 171Gb uncompressed). Visually they show significant variation in colour, resolution, and contrast.

**Figure 3.**
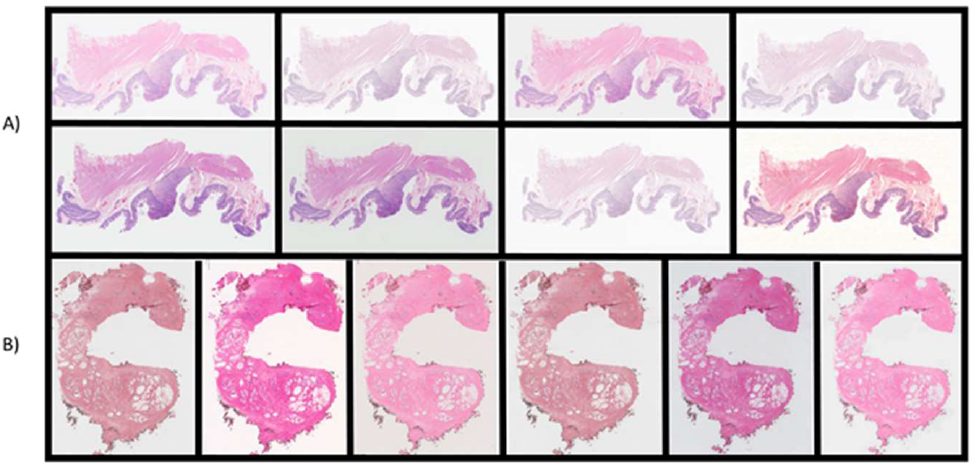
Slide replication across all digital pathology systems in the AI FORGE. A) A single 3”x1” slide scanned across 8 platforms (Leica Biosystems GT450 and AT2, Roche DP600, Hamamatsu S360MD and S20, 3DHistech P250 Flash, Akoya Bioscience PhenoImager, and Grundium Ocus). B) A single 3”x2” slide scanned across all 3”x2” capable scanners (Roche DP200; Hamamatsu S60 and S60v2MD, Olympus VS200, Objective Imaging Glissando, and OptraScan - OS-Ultra). The order of the images has been randomised from the listed order of platforms.

The FORGE has a total slide capacity of 3087 with all systems fully loaded, however the capacity to continuously load several systems allows the AI FORGE to scan approximately 4000 slides per day, which generates up to 7Tb of image data per day. Brightfield WSI scanning can be accomplished across all 15 systems, with fluorescence WSI capability across 3 systems (Olympus VS200, Hamamatsu S60, and Akoya PhenoImager HT). In addition, WSI Z-stacking can be delivered on 8 systems (S360MD, S60, S60v2, S20 Hamamatsu’s, 3DHistech P250 Flash, Olympus VS200, OptraScan OS-Ultra, and Objective Imaging Glissando).

### Quality control and quality assurance

Quality control (QC) and quality assurance (QA) processes are central to the production of high-quality images for diagnosis and AI training. NPIC have established QC processes which utilise off the shelf and in-house software for the review of digital images.

Manual evaluation of every image produced by the AI FORGE is undertaken by a suitably trained digital pathology team, working to established standard operating procedures (SOPs) ensuring consistency in the evaluation process. Image QC failure rate can be captured for every slide including categorical reasons for failure prior to rescanning to ensure images produced for AI training are free from errors which may influence AI algorithm application. Additionally, these failed images represent a useful source of AI training data which will be useful in training AI quality evaluation algorithms. Examples of artifact categories are shown in figure 4, including but not limited to, banding, debris, excess mountant, focus, marker pen, as well as multiple artifact failure possibilities. In addition to digital artefacts, the possibility of capturing pre-digitisation issues also exists, for example: tissue folds and tears, air bubbles, and microtomy chatter.

**Figure 4.**
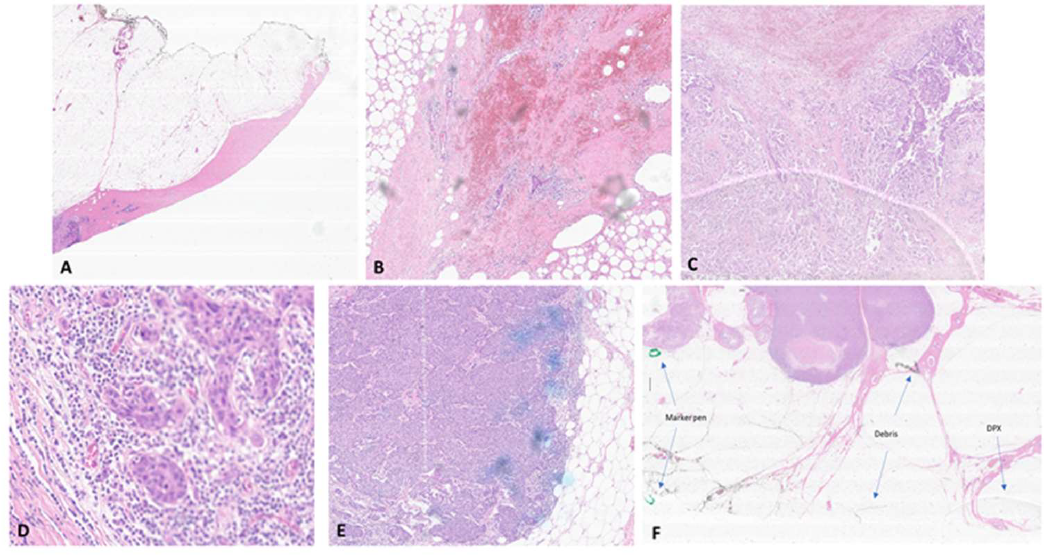
Common artifact categories manually identified during the image QC process. A) Banding. B) Debris. C) Excess mountant. D) Focus. E) Marker pen. F) Multiple artifacts (marker pen, debris, and DPX mountant indicated by the blue arrows).

Alongside the QC of scanned images is the QA process within the AI FORGE. In addition to the annual scanner servicing performed by manufacturers, daily QA assessments are undertaken for each scanner, in line with their individual use, care, and maintenance. Scanner settings are confirmed and recorded, inclusive of the international colour consortium (ICC) profile settings of each scanner (https://www.color.org/), image compression type, and compression quality applied. Adherence to the requirements of ISO:15189 enables us to ensure the accuracy and reliability of our processes and demonstrate the quality of service. Established processes enable secure specimen receipt, handling, transport, tracking, storage, and reporting. Furthermore, incident reporting, non-conformance and corrective actions are recorded within an auditable quality management system, inclusive of the training and competencies of all staff in the AI FORGE. Additionally, daily QC is facilitated by scanning of control slides. These include standard histological controls as well as custom histological phantoms or test objects developed in-house [12-14]. These control slides facilitate an objective ground truth enabling longitudinal image comparison.

### Facility

The laboratory space, originally a standard biological wet lab, was refurbished to create a dedicated digital pathology facility. A physical access control system grants access to employees by electronically authenticating their personal identity verification credentials via near-field communication access cards. Additional door codes are required for access to laboratory areas, requiring input of a secure code by keypad.

The laboratory space measures 46.75m^2^ (5.5m x 8.5m). Physical glass slide storage with capacity for up to 500,000 slides is present in the form of short- and long-term storage areas, a quick-access histology storage cabinet with capacity for 150,000 1”x3” slides, and modular archival storage capacity of 350,000 1”X3” slides.

To provide a secure housing for the scanners solid, 5cm thick, bespoke benching was installed. The benching was affixed to the concrete floor substructure and bolted to the walls. The depth of the benching was 850cm, to accommodate the footprint of each scanner. Powder-coated steel reinforced supports (5cm x 5cm) were installed to minimise movement and reduce potential vibration.

Appropriate air-conditioning, with redundancy was installed to maintain a consistent temperature of 20°C in the scanner room. Scanner heat-output for 15 scanners was calculated from the manufacturer’s specifications at 13,050 BTUs/per hour (3.82KW). The power required to cool a 46.75m2 room was calculated at 20,473 BTUs/per hour (6KW). Two wall mounted Mitsubishi Electric air conditioning units were installed capable of delivering a combined cooling power of 10KW (5KW each), offering over-capacity and redundancy in room cooling.

Sufficient electrical power needs were identified by referring to the manufacture’s specifications for all 15 scanners - 10.8KW was required for the operation of all systems. The power capabilities of the room were more than sufficient to support this, at 96.72KW. The electrical supply for each scanner instrument is supported by uninterrupted power supplies (UPS) rated at 2200VA with power conditioning that protects connected loads from electrical surges and spikes, lightning and other power disturbances. UPS will allow a scanner to run for an additional 20-30 minutes, providing the time to safely power off the system if power is interrupted mid-scan.

The physical workflow of the facility was also considered in the design process. Dedicated space for the reception of glass slides was created. Here all slides are receipted, booked-in, given a project and slide number, deidentified, and cleaned with 70% ethanol prior to scanning. Following the preparation of the slide, a process of multi-scanner digitisation can be undertaken. Batches of slide can be quickly moved between scanners, positioned less than 1m from one another.

All 15 digital pathology systems were installed in the room - figure 5 shows the current NPIC AI FORGE as of September 2023.

**Figure 5.**
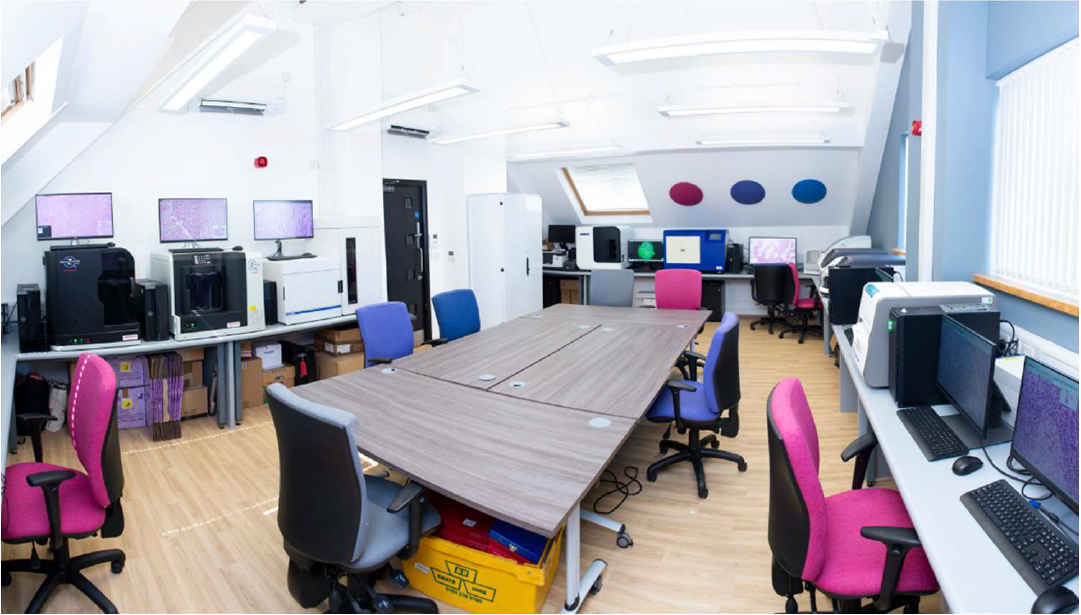
NPIC AI FORGE post installation image. Image shows the FORGE layout with a central island of tables for the receipt, preparation, and post scanning activity of glass slides with a quick access slide cabinet less than 1 meter away. Scanners are spaced with sufficient access allowance. Operator workstations are positioned throughout the facility and all monitors are high grade, manufacturer provisioned, or NPIC sourced medical grade displays. Sufficient lighting and air-conditioning capacity can also be seen.

### Training

Following the installation of the WSI systems, the NPIC scanning team of 8 underwent certified advanced training from each scanner manufacturer in the use, care and maintenance of each piece of equipment. Annual competency assessments are undertaken to ensure knowledge and training is current. Training and competency records for the team are recorded in line with ISO 15189 standards. In addition, every team member is trained in health and safety, fire safety, information governance, and data security.

### Power, networking, and data storage

The NPIC AI FORGE research storage is hosted by a technical partner (Exponential-e), who provide specialist connectivity, cloud and unified communications solutions (https://www.exponential-e.com/). The digital pathology WSI platforms are connected to the hospital network with a 10Gb/s connection, which in turn is connected by two 10Gb/s lines to the data centre. All 15 scanners are capable of producing 270MB of data per second.

Our data centre processes and stores images using Dell EMC PowerScale (https://www.dell.com/). Our fully managed platform spans two 2-tier data centres and provides high speed connectivity to partners on the NPIC programme. Within the datacentre environment the NPIC AI FORGE has 3 petabytes of dedicated expandable SSD storage in the form of 45TB of high-performance flash storage, with the remainder being Isilon storage.

The current data storage capacity for research is capable of hosting approximately 2 million images (average image size 1.5Gb). Our projected annual generation of images is currently in the region of 250,000 images. The research storage adheres to ISO standards for data storage (ISO 27001, ISO 9001 and ISO 14001), disaster recovery (ISO 27001), business continuity (ISO 22301), security (ISO 27017), ensuring compliance, security and reliability.

### Research image management system

Images generated from the 15 scanners are securely stored within our externally hosted data centre. For the Image ingestion, management, viewing, annotation and analysis of the images Halo Link (Indica Labs, UK) was provisioned as a research image management system. Halo Link enables the AI FORGE to automatically manage, share and analyse images securely through a web-based interface.

NPICs 3 petabytes of data storage is serviced by a dedicated expandable GPU processing resource for image analysis, including the training and validation of AI. Utilising multiple NVidia A40, A5000 and A6000 graphics cards. The NPIC AI FORGE’s current processing capacity has a combined throughput of 2 petaFLOPS.

Additionally, the facility is equipped with PC workstations including high resolution and medical-grade displays for the viewing, image analysis, and QC of images generated. The minimum specifications of the workstations are as follows: i5-11500 6 core 4.6GHz processor, 16gb memory, 512GB SSD, 1TB HDD, and a 8GB AMD Radeon GPU, with 4 display ports powering Jusha C620 Diagnostic 6MP medical grade displays (http://www.jusha.com.cn/).

## Discussion

This manuscript presents the establishment and operation of the NPIC AI FORGE, a unique multi-scanner facility within an NHS hospital, aimed at (a) scanner evaluation and (b) facilitating the replication of image cohorts for the training and development of AI algorithms across multiple WSI pathology systems. The NPIC AI FORGE currently comprises a collection of 15 high-throughput digital pathology WSI scanners from 9 different manufacturers.

We believe the AI FORGE comprises the broadest collection of clinical grade digital pathology WSI systems in a single environment. Our aim in the creation of the AI FORGE was to represent a significant proportion of the clinical market of digital pathology systems in a single environment to facilities the replication of image cohorts for the training and development of AI algorithms. The capability of the facility has the potential to improve diagnostic accuracy and efficiency of AI, as well as address the limitations and challenges associated with pathology-based AI.

We did not set out to create a facility capable of digitally scanning tens of thousands of glass slides per day. Facilities exist that are able to facilitate larger capacity scanning projects, utilising several single manufacturer systems. However, to date, the broad capabilities demonstrated by the AI FORGE have not been brought together in a single accessible facility, to our knowledge.

The ability to acquire and analyse high-resolution digital images of entire pathology slides across a range of WSI scanners has the potential to generate AI training data beyond the current industry standard. AI tools built on training data from diverse images may provide further algorithmic robustness and resilience enabling a higher degree of generalisability from classically trained tools. However, diversity from different hospital locations alone does not increase algorithm generalisability by default, and may be less data-efficient than a targeted replication of smaller datasets on multiple platforms. Evaluation and training of AI models from diverse sites have also been shown to demonstrated AI model overfitting and bias. It was recently shown that an AI model trained on The Cancer Genome Atlas (TCGA) image data, collected from diverse sites, resulted in clear batch signatures which lead to a biased accuracy in the predictive capabilities of the model [15]. The authors note that consideration for other factors alongside image diversity should be evaluated, for example image compression quality and stain variation. The authors suggest that stain normalization may remove some variation and image augmentation could reduce differences in colour. Though the authors attempted to minimised image resolution disparity across the images by sampling only a fixed pixel to μm ratio, they did not assess compression directly. But data from elsewhere confirms that JPEG quality has a strong negative effect on the performance of AI on tasks using TCGA images [16]. These factors as well as others such as application of ICC profiles, choice of scanner, and sample acquisition methodology can all be closely controlled within the AI FORGE.

It is important to acknowledge that higher volumes of training data alone may not be sufficient to overcome the brittleness of AI and consideration for the diversity of the source data must be considered in the training of AI, as mentioned above. Studies training AI on many thousands of images reveal that even high volumes training data may suffer from overfitting and a lack of generalisability. For example, a comparative study in dermatopathology of over 130 machine-learning algorithms trained on over 10,000 dermatological images reported that the superior performance seen was likely due to the overfitting of the training data. The authors acknowledge that algorithm overfitting was a limitation that led to a lack of generalisability on images beyond the training set [17, 18]. The addition of more data may also introduce more complexity and models can be susceptible to shortcut learning based on confounding variables such as scanner type or laboratory origin [19].

We recognise that the provision of images alone from multiple digital pathology scanners for AI training will likely not by itself solve the issue of overfitting and the poor generalisability seen with some AI tools. This is clear from the development of AI in radiology where poor generalisability seen on images from various imaging equipment is likely due to limited or in some cases absent training data from respective imaging equipment sources [20, 21]. A retrospective study classifying chest radiographs as normal or abnormal obtained over 200,000 chest images; however, the AI algorithm showed little improved performance after training on 20,000 radiographs [22].

This has led to many AI vendors in radiology restricting the application of their AI solutions to images generated by specific equipment, which leads to vendor lock-in [23]. The ethical and practical barriers to obtaining multiple images of the same patient across several radiological modalities for the use in AI training data does not exist in pathology. Multiple rounds of imaging of pathology slides across several pieces of equipment has no impact on the patient, unlike the increased risk from obtaining repeated CT scans from a single patent, for example. This reality provides an opportunity in pathology to develop broader and more diverse training data, while minimising effort in data gathering.

As a facility scanning clinical slides from patients, it is important in designing such a facility that appropriate measures are taken to adhere to data protection regulations and gain ethical approval for data use as well as patient and public involvement in the use of patient data for research.

While the NPIC AI FORGE represents a step forward in the integration of AI and digital pathology, there are still challenges that need to be addressed. Version control and iterative improvements in AI algorithms are necessary to ensure their optimal performance. Regular validation and testing of AI models in real-world clinical scenarios will be crucial in building confidence in AI-based pathology and its integration into routine practice.

## Conclusion

The NPIC AI FORGE has been established to address the challenges and limitations of AI-based pathology by providing access to large, diverse, and high-quality datasets from multiple WSI systems. The establishment of this facility significantly contributes to the advancement of AI algorithms in digital pathology, potentially leading to improved diagnostic accuracy, workflow efficiency, and ultimately improved patient care. As NPIC continues to refine the AI FORGE, it is anticipated that this facility will serve as an exemplar for other healthcare institutions globally, improving the quality and efficiency of data gathering for AI and hence accelerating the adoption of AI in pathology.

## Data Availability Statement

Data within this study are available from the corresponding author, upon reasonable request.

## References

1. Patel A, Balis UGJ, Cheng J, Li Z, Lujan G, McClintock DS, et al. Contemporary Whole Slide Imaging Devices and Their Applications within the Modern Pathology Department: A Selected Hardware Review. (2229–5089 (Print)).

2. Magee DR, Treanor D, Chomphuwiset P, Quirke P, editors. Context Aware Colour Classification in Digital Microscopy 2011.

3. Clarke EL, Treanor D. Colour in digital pathology: a review. Histopathology. 2017;70(2):153–63.

4. Magee D, Treanor D, Crellin D, Shires M, Smith K, Mohee K, et al., editors. Colour normalisation in digital histopathology images2009: Daniel Elson London.

5. Clarke EL, Brettle D, Sykes A, Wright A, Boden A, Treanor D. Development and Evaluation of a Novel Point-of-Use Quality Assurance Tool for Digital Pathology. (1543–2165 (Electronic)).

6. Allan G, Alex W, Pete J, Mike H, Darren T. Quantification of histochemical stains using whole slide imaging: development of a method and demonstration of its usefulness in laboratory quality control. Journal of Clinical Pathology. 2015;68(3):192.

7. Williams BA-O, Knowles C, Treanor D. Maintaining quality diagnosis with digital pathology: a practical guide to ISO 15189 accreditation. (1472–4146 (Electronic)).

8. Wright AI, Dunn CM, Hale M, Hutchins GGA, Treanor DE. The Effect of Quality Control on Accuracy of Digital Pathology Image Analysis. IEEE Journal of Biomedical and Health Informatics. 2021;25(2):307–14.

9. Treanor D, & Williams BJ B.. The Leeds Guide to Digital Pathology.. 2018.

10. von Eschenbach WJ. Transparency and the Black Box Problem: Why We Do Not Trust AI. Philosophy & Technology. 2021;34(4):1607–22.

11. Europe H. Digital Pathology Solutions

12. Clarke EL, Revie C, Brettle D, Shires M, Jackson P, Cochrane R, et al. Development of a novel tissue-mimicking color calibration slide for digital microscopy. Color Research & Application. 2018;43(2):184–97.

13. Hayley Pye DB, Danny Kaye, Catriona M. Dunn, Matthew P. Humphries, M. P., and Darren Treanor. A First Look at Scanner Introduced Variation in Contrast, Resolution, and Colour Across 5 Different Models of Whole Slide Imaging (WSI) Scanner. 2022.

14. Catriona Dunn DB, Martin Cockroft, Elizabeth Keating, Craig Revie, and Darren Treanor. The use of a biopolymer film for quantitative H&E stain assessment and quality control in pathology. 2022.

15. Howard FM, Dolezal J, Kochanny S, Schulte J, Chen H, Heij L, et al. The impact of site-specific digital histology signatures on deep learning model accuracy and bias. Nature Communications. 2021;12(1):4423.

16. Komura D, Ishikawa S. Machine Learning Methods for Histopathological Image Analysis. (2001–0370 (Print)).

17. Tschandl P, Codella N, Akay BN, Argenziano G, Braun RP, Cabo H, et al. Comparison of the accuracy of human readers versus machine-learning algorithms for pigmented skin lesion classification: an open, web-based, international, diagnostic study. (1474–5488 (Electronic)).

18. Kuo KM, Talley PC, Chang CS. The accuracy of artificial intelligence used for non-melanoma skin cancer diagnoses: a meta-analysis. (1472–6947 (Electronic)).

19. Schmitt MA-O, Maron RA-OX, Hekler AA-O, Stenzinger AA-OX, Hauschild AA-O, Weichenthal MA-O, et al. Hidden Variables in Deep Learning Digital Pathology and Their Potential to Cause Batch Effects: Prediction Model Study. (1438–8871 (Electronic)).

20. Bluemke DA-O, Moy LA-O, Bredella MA-O, Ertl-Wagner BA-O, Fowler KA-O, Goh VA-O, et al. Assessing Radiology Research on Artificial Intelligence: A Brief Guide for Authors, Reviewers, and Readers-From the Radiology Editorial Board. (1527–1315 (Electronic)).

21. Putha P, Tadepalli M, Reddy B, Raj T, Chiramal JA, Govil S, et al. Can artificial intelligence reliably report chest x-rays?: Radiologist validation of an algorithm trained on 2.3 million x-rays. arXiv preprint arXiv:180707455. 2018.

22. Dunnmon JA-O, Yi D, Langlotz CP, Ré C, Rubin DA-O, Lungren MP. Assessment of Convolutional Neural Networks for Automated Classification of Chest Radiographs. (1527–1315 (Electronic)).

23. Leiner TA-O, Bennink E, Mol CP, Kuijf HJ, Veldhuis WB. Bringing AI to the clinic: blueprint for a vendor-neutral AI deployment infrastructure. (1869–4101 (Print)).

